# Quantitative evaluation of *MGMT* promoter methylation in FFPE and fresh frozen brain tumor samples by EpiDirect MGMT Methylation qPCR Assay compared to pyrosequencing: correlation and corresponding cutoffs

**DOI:** 10.64898/2026.11.09.26367351

**Authors:** Michael Børgesen, Maria Vad Jakobsen, Linea Cecilie Melchior, Nanna Vesterskov Jensen, Kamilla Bendixen, Iben Daugaard, Hanne Poulsen, Morten Grauslund

## Abstract

*MGMT* promoter (*MGMTp*) methylation is a clinically important epigenetic biomarker that guides therapeutic decision-making in patients with glioblastoma and related brain malignancies. Pyrosequencing is a widely used method for analysis of *MGMTp* methylation in molecular pathology laboratories which relies on chemical conversion of DNA prior to amplification by polymerase chain reaction (PCR) and subsequent sequencing. The EpiDirect^®^ technology represents a novel analytical approach that enables direct, quantitative assessment of DNA methylation by qPCR using native, unmodified DNA. Here we compared the EpiDirect^®^ MGMT Methylation qPCR Assay to *MGMTp* pyrosequencing using brain tumor samples to evaluate the correlation between the two methods. We show that there is a clear correlation across two different clinical cohorts involving both formalin-fixed paraffin-embedded (FFPE) and fresh frozen samples and show that the EpiDirect^®^ MGMT Methylation qPCR Assay is a valid alternative to pyrosequencing for quantitative assessment of *MGMTp* methylation in brain cancer samples.

## Introduction

Epigenetic modifications are increasingly used as biomarkers in precision medicine. In cancer, gene silencing by promoter DNA methylation is not only implicated in initiation of disease but also used to assist treatment selection^1–3^. In glioblastoma and related gliomas, DNA methylation of the promoter region of the *MGMT* gene (*MGMTp*) encoding the DNA repair enzyme O-6-methylguanine-DNA methyltransferase is a prognostic biomarker and a predicter of alkylating agent treatment efficiency^4–11^. Thus, overall survival of patients with methylated *MGMTp* is significantly higher when treated with radiotherapy plus the alkylating agent temozolomide compared to radiotherapy alone^9,12,13^. Furthermore, there is a direct positive correlation between the extent of *MGMTp* methylation and the overall survival of patients treated with temozolomide^14,15^. Evaluation of *MGMTp* methylation is therefore recommended as part of the molecular characterization of high-grade gliomas^16,17^.

The human *MGMT* gene is located on chromosome 10 and contains a CpG island within its promoter consisting of 98 CpG sites between (GRCh38) positions 129,466,685 and 129,467,446 (**Figure 1**)^15,18,19^. In glioblastoma patients, the *MGMTp* contains differently methylated regions with different clinical significance^15,19–22^. Especially one differentially methylated region (CpG 73-90, DMR-2^19^) shows high correlation to the general methylation status of the *MGMTp* as well as to the overall survival of patients treated with temozolomide^15,22^.

**Figure 1.**
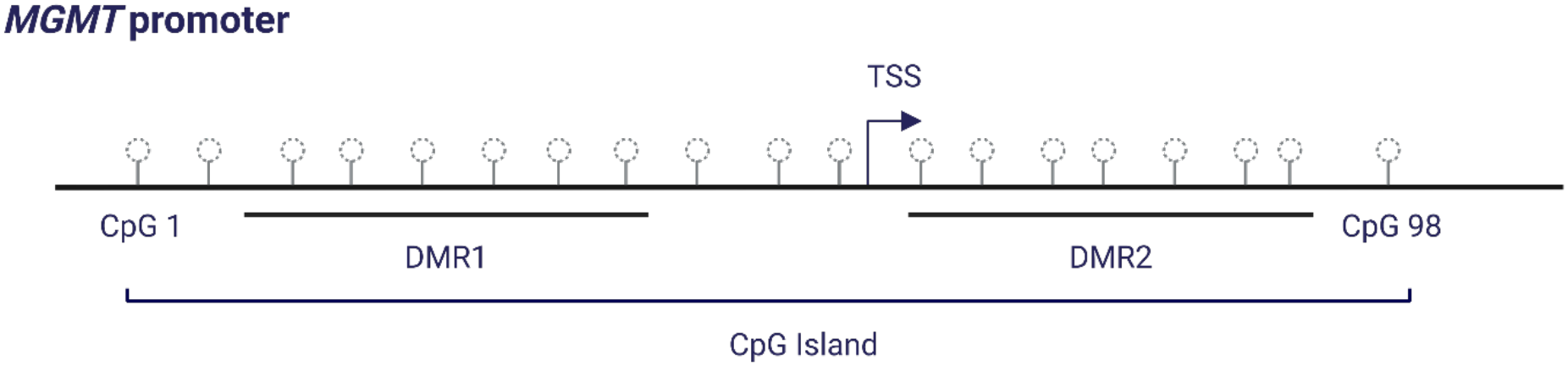
The human *MGMT* promoter contains a CpG island at chromosome 10 position 129,466,685 to 129,467,446 (GRCh38) consisting of 98 CpG sites and two differentially regulated regions (DMR1 and DMR2)^19^ near the transcription start site (TSS).

Current methods for evaluating *MGMTp* methylation mainly involve chemical or enzymatic conversion of the purified DNA prior to selective amplification by PCR of the target region of interest. In most cases, chemical conversion of DNA is performed using sodium bisulfite, which converts unmethylated cytosine nucleobases to uracils while not affecting methylated cytosines^23^. The converted uracils are subsequently replaced by thymine nucleobases in the replicated DNA strands during the PCR step, and this change in sequence can be assessed by either pyrosequencing or methyl-specific PCR methods^23,24^. Other methods for evaluation of *MGMTp* methylation not relying on chemical conversion of DNA include methylation-sensitive digestion by restriction enzymes followed by ligation-dependent probe amplification (MS-MLPA)^25^ and nanopore sequencing^26^. However, both chemical and enzymatic conversion of DNA as well as sequencing procedures are time- and resource-consuming, and in addition, chemical conversion leads to DNA degradation, which appears to be methylation-dependent and may thus lead to overrepresentation of the methylated DNA^27^.

The *therascreen*^*®*^ MGMT Pyro Kit (Qiagen, described here as the *therascreen*^*®*^ MGMT Kit) is a widely used pyrosequencing method to evaluate *MGMTp* methylation status of brain tumor samples. The *therascreen*^*®*^ MGMT Kit targets four CpG sites (CpG 76-79) in the DMR2 of the *MGMTp* (**Figure 2**), and, as with most other current methods for evaluating DNA methylation, purified DNA needs to be bisulfite-treated prior to analysis by the *therascreen*^*®*^ MGMT Kit method.

**Figure 2.**
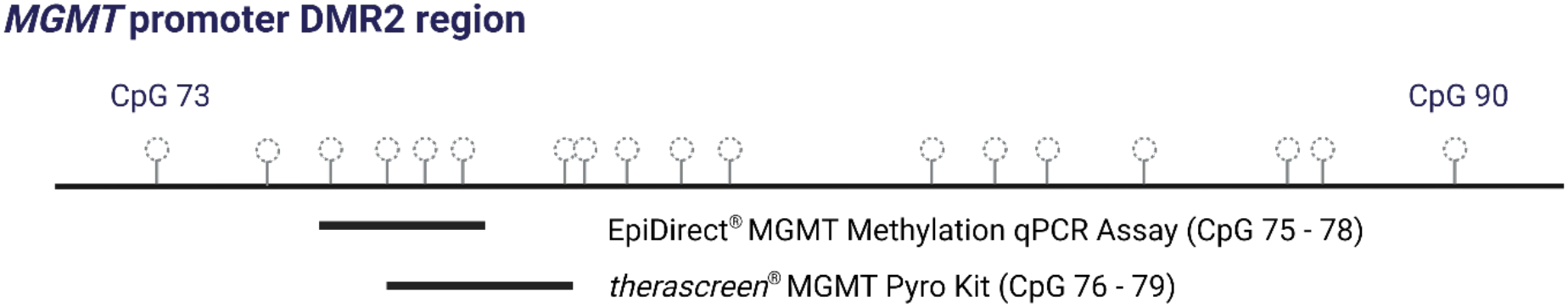
The EpiPrimer™ in the EpiDirect^®^ MGMT Assay is placed in the differentially methylated region 2 (DMR2, GRCh38 chr10; 129,467,232-129,467,363) of the *MGMT* promoter^19^ and covers CpG 75-79^28^. DMR2 methylation has been shown to correlate strongly with overall *MGMT* promoter methylation status as well as with temozolomide treatment efficiency^15,18,22^.

The EpiDirect^®^ MGMT Methylation qPCR Assay (“EpiDirect MGMT Assay”, PentaBase A/S) is a novel method for direct quantification of *MGMTp* methylation in untreated DNA, as recently described^28^. The method is based on direct selective amplification of methylated *MGMTp* DNA using an EpiPrimer™, which contains hydrophobic base analogues^29–32^ giving the EpiPrimer™ its unique affinity for methylated DNA. The EpiDirect^®^ Assay also evaluates the methylation status of four CpG sites in the DMR2 of the *MGMTp* (CpG 75-78), thus overlapping with three of the CpG sites evaluated by the *therascreen*^*®*^ MGMT Kit (**Figure 2**)^15,18,22^.

Motivated by the upcoming discontinuation of the *therascreen*^*®*^ MGMT Kit, we conducted both retrospective and prospective comparative studies between the EpiDirect^®^ MGMT Assay and the *therascreen*^*®*^ MGMT Kit, using various brain cancer sample types as well as both formalin-fixed, paraffin-embedded (FFPE) and fresh frozen samples. We also compared results obtained with the EpiDirect^®^ MGMT Assay when using either pre- defined algorithms or standard curve-derived algorithms for calculating sample methylation percentages. We demonstrate a convincing correlation between the results obtained with the EpiDirect^®^ MGMT Assay and those generated by the *therascreen*^*®*^ MGMT Kit. *MGMTp* methylation frequencies were generally lower with the EpiDirect^®^ MGMT Assay, and when comparing results to those obtained with the *therascreen*^*®*^ MGMT Kit with a 10% cutoff, a 5% and 7% methylation cutoff for the EpiDirect^®^ MGMT Assay yielded similar results when analyzing FFPE samples used in Cohort 1 and fresh frozen samples used in Cohort 2, respectively.

## Methods

### *MGMT* methylation assays

The EpiDirect^®^ MGMT Methylation qPCR Assay (“EpiDirect^*®*^ MGMT Assay”, PentaBase A/S, Odense, Denmark Ref. No. 8300) was used according to the Instructions for Use (IFU) version 2.8. The *therascreen*^*®*^ MGMT Pyro Kit (“*therascreen*^*®*^ MGMT Kit”, Qiagen, Hilden, Germany, Cat no. 971061) was used according to IFU version 1 with the PyroMark Q24 instrument (Qiagen) and the PyroMark Q24 Software, v.2.0.8.

#### Cohort 1

Cohort 1 was used in a prospective study involving brain tumor samples collected in Denmark by Aarhus University Hospital, Aarhus, Denmark during the fall of 2024 and the spring of 2025. Patients were between 27 and 86 years of age diagnosed with either glioblastoma, oligodendroglioma, glioma, or a brain tumor with an unspecific alteration (**Supplementary Table 1**). Inclusion criteria were histologically proven brain tumor samples containing at least 20% tumor tissue with available leftover tumor tissue or DNA for EpiDirect^®^ MGMT Assay evaluation. DNA was purified using the EZ1&2 DNA FFPE Kit (Qiagen Cat no. 954404) and DNA concentration was determined by the Qubit fluorometer (Qiagen) according to the manufacturer&’s instructions. Exclusion criteria were purified sample DNA concentrations below 3.0 ng/&µL and more concentrated samples were diluted to 3.0 ng/µL before analysis by the EpiDirect^®^ MGMT Assay. Before analysis by the *therascreen*^*®*^ MGMT Kit, purified DNA was bisulfite converted using the MethylEdge™ Bisulfite Conversion System (Promega Cat no. N1301) according to the manufacturer’s instructions. The EpiDirect^®^ MGMT Assay was run on both a QuantStudio™ 5 (Applied Biosystems) and the BaseTyper™ 48.4 (PentaBase A/S) Real-Time PCR System and was used in combination with the Methylated Human DNA Control Panel (PentaBase A/S, Ref. No. 5070) for the generation of DNA methylation standard curves including 3%, 5%, and 10% methylated samples in addition to the 25% methylated positive control. The slope and y-axis intercept of the resulting trendline were used to calculate run-specific sample methylation percentages in addition to calculations based on the pre-defined algorithms described in the EpiDirect^®^ MGMT Assay IFU version 2.8. To determine the impact of decreasing DNA input on methylation percentages determined by the EpiDirect^®^ MGMT Assay, selected patient DNA samples were diluted in nuclease-free water down to 0.5 ng/µL.

#### Cohort 2

Cohort 2 was collected at Rigshospitalet, Copenhagen, Denmark, between February and December 2025 as fresh frozen tissue samples for nanopore sequencing. Patients were between 18 and 89 years of age diagnosed with either glioblastoma, astrocytoma, oligodendroglioma, or not otherwise specified (NOS) glioma (**Supplementary Table 2**). DNA was purified using the Maxwell RSC Blood DNA kit (Promega, Madison, WI, No. AS1400) on a Maxwell RSC instrument. DNA was diluted to 2 ng/µL before MGMT testing by the EpiDirect^®^ MGMT Assay using a QuantStudio™ 5 Real-Time PCR System and using the pre-defined algorithm for calculating sample methylation percentage as described in EpiDirect^®^ MGMT Assay IFU version 2.8.

### Data analysis and statistical methods

#### EpiDirect^®^ MGMT Assay

Data analysis was performed according to the EpiDirect^®^ MGMT Methylation qPCR Assay IFU version 2.8 as well as the Methylated Human MGMT and MLH1 DNA Control Panel (ref. no. 5070) IFU version 1.0. Baselining was performed according to the manufacturer’s instructions, and the threshold was set in each run at 10% of the mean maximum relative fluorescence units (RFU) level of the positive control duplicate reactions^a^. For each sample and control, the Ct values for both the Methyl-specific Assay and the Internal Reference Assay were determined, and the delta Ct was calculated by subtracting the Ct value of the Internal Reference Assay from the Ct value of the Methyl-specific Assay (**Equation 1**):

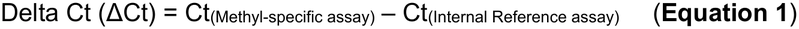

The delta Ct values were subsequently used to calculate the sample and control *MGMTp* methylation percentage using the predefined algorithm described in the EpiDirect^®^ MGMT Assay IFU version 2.8 (**Equation 2**):

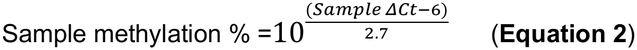

In cohort 1, sample methylation percentages were also calculated using the algorithms generated by plotting the delta Ct values of the 3%, 5%, 10% standards and positive control (25%) as a function of the common logarithm (log_(10)_) to the methylation percentage. The resulting equation describing the linear regression (**Equation 3)** was used to calculate the methylation percentages of the samples included in each run (**Equation 4**).

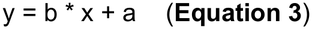

where y is the ΔCt value, x is the common logarithm to the methylation percentage, a is the y-axis intercept, and b is the slope of the trendline.

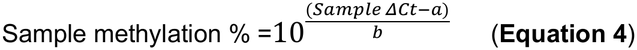

### therascreen^®^ MGMT Kit

The methylation percentages of each of the 4 included CpG sites were determined, and the average methylation percentage was calculated. A 10% average methylation percentage was used as the cutoff between methylated and unmethylated samples.

### ROC curves

ROC curve analysis with Youden’s Index was performed using RStudio (2025.05.1 Build 513) with R version 4.5.1 to calculate the optimal EpiDirect^®^ MGMT Assay methylation percentage cutoff that corresponds to a 10% methylation cutoff used with the *therascreen*^*®*^ MGMT Kit as well as the EpiDirect^®^ MGMT Assay methylation percentage cutoff that leads to a similar negative (unmethylated) percent agreement (NPA) and positive (methylated) percent agreement (PPA) (**Supplementary Data 1 and Supplementary Data 2**).

### Ethical considerations

This study was based on leftover samples that the study sites have permission to use for evaluation of diagnostic assays. The results obtained with the EpiDirect^®^ MGMT Methylation qPCR Assay were only used for assay evaluation and not for diagnostic purposes. Thus, the original diagnosis and treatment of the individuals from whom the samples originated were not changed based on this study.

## Results

### Cohort 1

#### Analysis of purified DNA from FFPE samples

A total of 45 prospective brain tumor samples as well as two external quality assessment (EQA) samples were analyzed during the fall of 2024 and the spring of 2025 in parallel by the *therascreen*^*®*^ MGMT Kit and the EpiDirect^®^ MGMT Assay (**Supplementary Table 1)**. The EpiDirect^®^ MGMT Assay was run on both a QuantStudio™ 5 (Applied Biosystems) and a BaseTyper™ 48.4 (PentaBase A/S) Real-Time PCR System. *MGMTp* methylation percentages were calculated using both predefined algorithms as well as by including a standard curve in each run. Standard curves were generated by plotting the delta Ct values (Ct value obtained with the Methyl-specific Assay subtracted the Ct value obtained with the Internal Reference Assay) of the controls as a function of the common logarithm to their *MGMTp* methylation percentages, 3%, 5%, 10%, and 25% (**Supplementary Figure 1**). The methylation percentages of the individual samples were calculated from the included standard curve in the same run.

EpiDirect^®^ and pyrosequencing results show a fairly high degree of correlation for the *MGMTp* methylation percentages with an R^2^ of 0.81 and 0.78 when using the EpiDirect^®^ MGMT Assay on the QuantStudio™ 5 and the BaseTyper™ 48.4 Real-Time PCR System, respectively (**Figure 3**).

**Figure 3.**
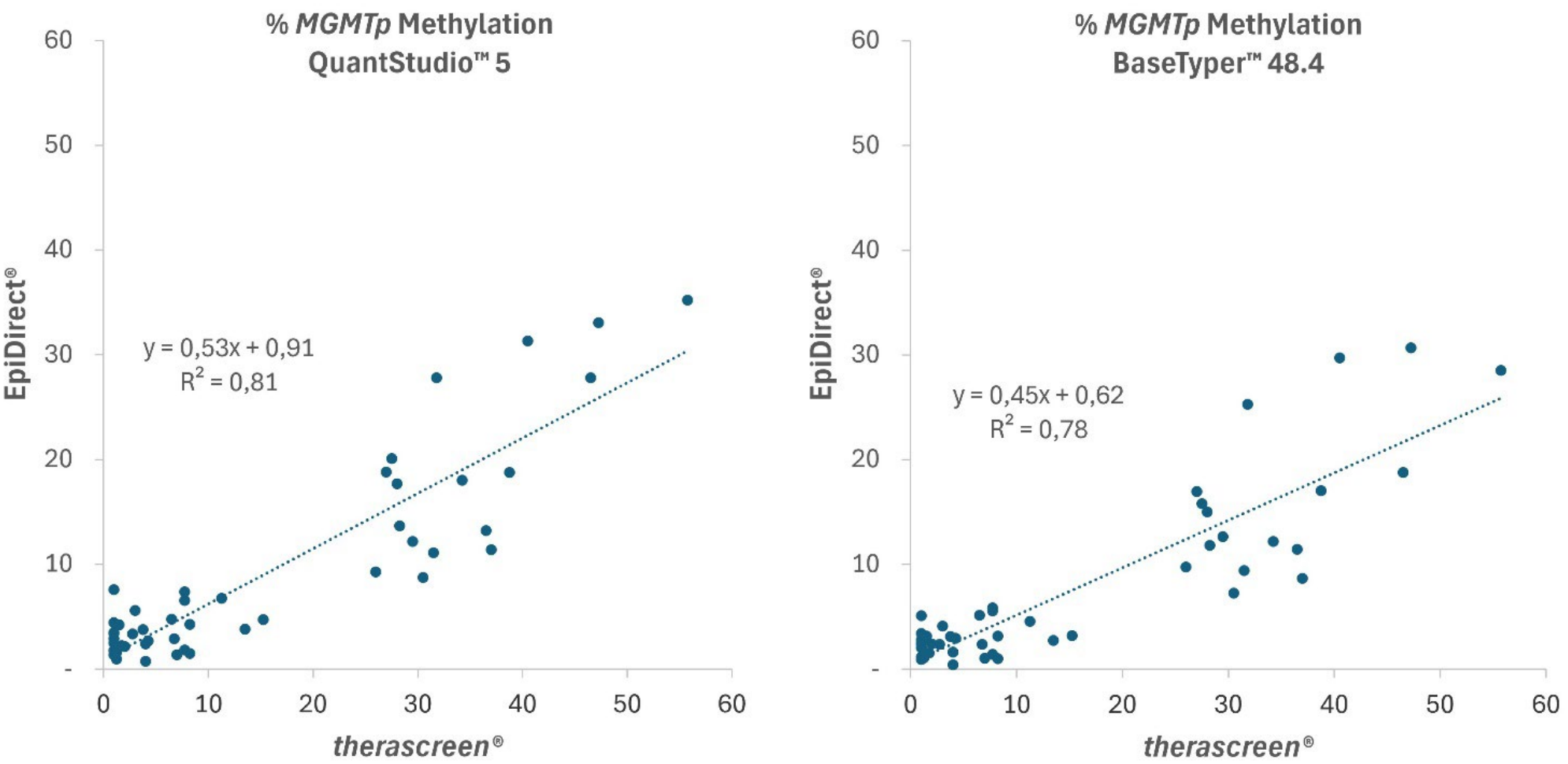
Cohort 1 *MGMT* promoter (*MGMTp*) methylation frequencies as determined by the EpiDirect^®^ MGMT Assay plotted against methylation frequencies of the same samples determined with the *therascreen*^*®*^ MGMT Kit. The EpiDirect^®^ MGMT Assay was used with the QuantStudio™ 5 (left) and the BaseTyper™ 48.4 real-time PCR systems (right). *MGMTp* methylation frequencies were calculated using the algorithms generated by including a standard curve in each run.

On both real-time PCR systems, the determined sample methylation percentages were generally lower when using the EpiDirect^®^ MGMT Assay compared to the *therascreen*^*®*^ MGMT Kit. Thus, linear regression slopes were 0.51 and 0.45 when using the EpiDirect^®^ MGMT Assay with the QuantStudio™ 5 and the BaseTyper™ 48.4 Real-Time PCR Systems, respectively. Similar results were found when using predefined algorithms for calculating methylation frequencies with the EpiDirect^®^ MGMT Assay (**Supplementary Figure 2**).

### Robustness across real-time PCR platforms

When directly comparing EpiDirect^®^ MGMT Assay results on the two different qPCR platforms, a high degree of correlation between the determined *MGMTp* methylation percentages was found when using both the IFU version 2.8 algorithms and the algorithms generated by including a standard curve in each run (**Figure 4**). While the slope is further from 1 and the R^2^ value is lower with the standard curve algorithms compared to the IFU algorithms, the opposite is true when excluding samples with methylation percentages above 20% that are far from the expected clinically relevant area, and the slope becomes very close to 1 (0.97) (**Supplementary Figure 3**).

**Figure 4.**
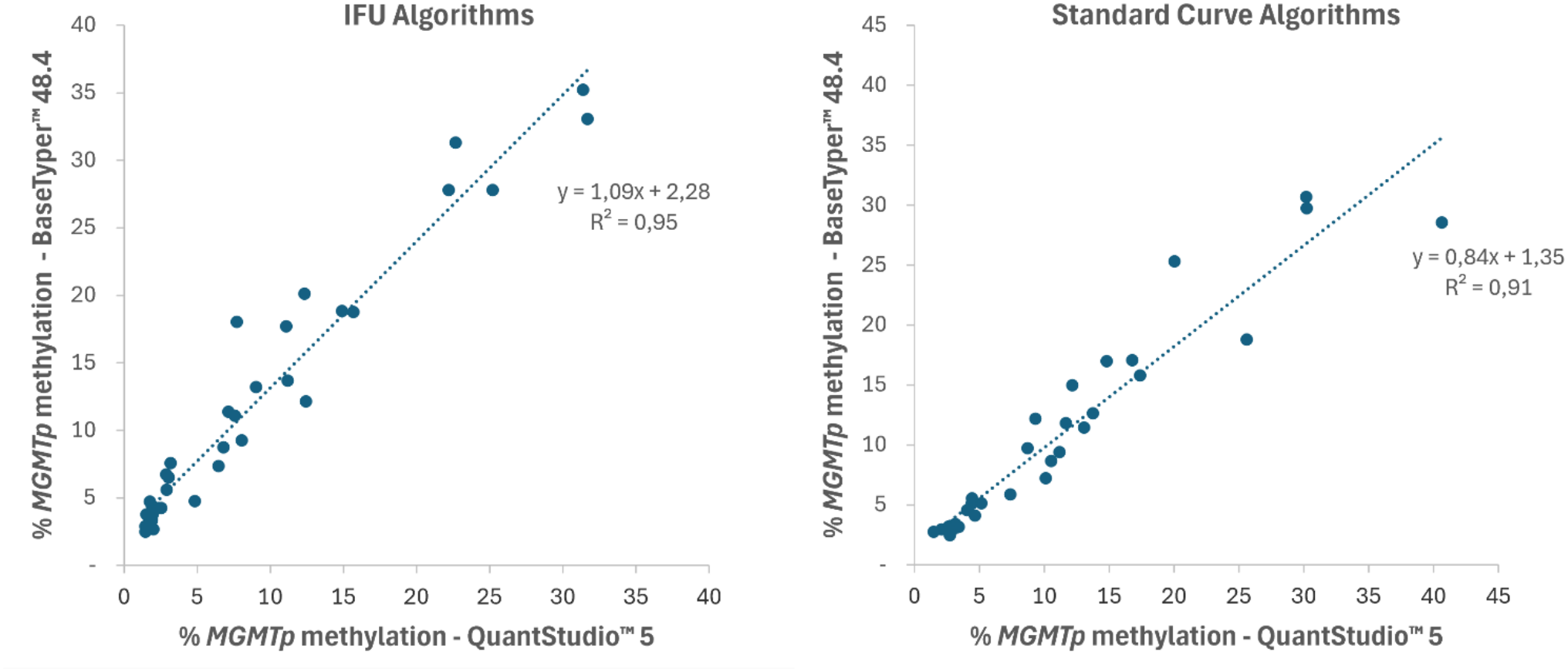
Correlation between *MGMT* promoter (*MGMTp*) methylation frequencies determined by the EpiDirect^®^ MGMT Assay when using the QuantStudio™ 5 and the BaseTyper™ 48.4 real-time PCR Systems in Cohort 1. Data are shown for *MGMTp* methylation frequencies calculated using the algorithms described in the EpiDirect^®^ MGMT Assay Instructions for Use (IFU) version 2.8 (left) and when including a standard curve in each run using the Methylated Human DNA Control panel (right). Samples below the limit of detection of the EpiDirect^®^ MGMT Assay are excluded.

#### Calculating Corresponding Cutoff for the EpiDirect^**®**^ MGMT Assay

A 10% average methylation cutoff across the four included CpG sites was used with the *therascreen*^*®*^ MGMT Kit for differentiating methylated samples from unmethylated samples clinically. To calculate the corresponding cutoff for the EpiDirect^®^ MGMT Assay, a receiver operating characteristic (ROC) curve analysis with Youden’s Index was performed (**Supplementary Data 1**). The resulting ROC curves and **corresponding** cutoffs for EpiDirect^®^ MGMT Assay are shown in **Supplementary Figure 4, Table 1** (QuantStudio™ 5) and **Table 2** (BaseTyper™ 48.4). The ROC curve analysis found optimal corresponding cutoffs for the EpiDirect^®^ MGMT Assay between 5.9% and 8.2% methylation across instruments and analysis methods (**Table 1** and **Table 2**). In all cases, these cutoffs resulted in 100% negative percent agreements (NPA), while the positive percent agreements (PPA) were lower.

**Table 1.** Receiver operating characteristic (ROC) curve analysis of Cohort 1 results for determination of corresponding cutoff when comparing EpiDirect^®^ MGMT Methylation qPCR Assay results on the QuantStudio™ 5 Real- Time PCR System with the *therascreen*^*®*^ MGMT Kit. A cutoff of 10% methylation was used for the *therascreen*^*®*^ MGMT Kit. * With Youden’s index. PPA, positive percent agreement (methylated samples). NPA, negative percent agreement (unmethylated samples). OPA, overall percent agreement. AUC, Area Under the Curve.

| **QuantStudio™ 5** |  |  |  |  |  |  |
| **Algorithm** | **Method** | **Cutoff (% methylation)** | **PPA** | **NPA** | **OPA** | **AUC** |
|---|---|---|---|---|---|---|
| **IFU algorithm** | Optimal cutoff* | 6.6 | 0.83 (15/18) | 1.00 (27/27) | 0.93 (42/45) | 0.95 |
|  | Similar PPA and NPA | 3.1 | 0.83 (15/18) | 0.89 (24/27) | 0.87 (39/45) |  |
| **Standard curve algorithm** | Optimal cutoff* | 8.1 | 0.83 (15/18) | 1.00 (27/27) | 0.93 (42/45) | 0.94 |
|  | Similar PPA and NPA | 4.6 | 0.83 (15/18) | 0.89 (24/27) | 0.87 (39/45) |  |
|  | Laboratory procedure | 5.0 | 0.83 (15/18) | 0.93 (25/27) | 0.89 (40/45) |  |

**Table 2.** Receiver operating characteristic (ROC) curve analysis of Cohort 1 results for determination of corresponding cutoff when comparing EpiDirect^®^ MGMT Methylation qPCR Assay results on the BaseTyper™ 48.4 Real-Time PCR System with the *therascreen*^*®*^ MGMT Kit. A cutoff of 10% methylation was used for the *therascreen*^*®*^ MGMT Kit. * With Youden’s index. PPA, positive percent agreement (methylated samples). NPA, negative percent agreement (unmethylated samples). OPA, overall percent agreement. AUC, Area Under the Curve.

| **BaseTyper™ 48.4** |  |  |  |  |  |  |
| **Algorithm** | **Method** | **Cutoff (% methylation)** | **PPA** | **NPA** | **OPA** | **AUC** |
|---|---|---|---|---|---|---|
| **IFU algorithm** | Optimal cutoff* | 8.2 | 0.83 (15/18) | 1.00 (27/27) | 0.93 (42/45) | 0.97 |
|  | Similar PPA and NPA | 6.1 | 0.89 (16/18) | 0.89 (24/27) | 0.89 (40/45) |  |
| **Standard curve algorithms** | Optimal cutoff* | 5.9 | 0.83 (15/18) | 1.00 (27/27) | 0.93 (42/45) | 0.96 |
|  | Similar PPA and NPA | 4.5 | 0.89 (16/18) | 0.85 (23/27) | 0.87 (39/45) |  |
|  | Laboratory procedure | 5.0 | 0.83 (15/18) | 0.85 (23/27) | 0.84 (38/45) |  |

We therefore also evaluated which *MGMT* methylation cutoffs would produce similar PPA and NPA values (method described in **Supplementary Data 2**). In this case, the cutoffs were between 3.1% and 6.1% when using IFU algorithms and between 4.5% and 4.6% when using standard curve algorithms, supporting the notion that the use of standard curves for calculating sample methylation percentages leads to more similar results across different real-time PCR platforms.

The clinical site decided to use a 5% methylation cutoff with the EpiDirect^®^ MGMT Assay and standard curve algorithms which leads to highly similar relative proportions of methylated vs. unmethylated samples when comparing EpiDirect^®^ MGMT Assay with *therascreen*^*®*^ MGMT Kit results (**Table 1** and **Table 2**). With standard curve-derived algorithms, this gave a PPA of 0.83 on both instruments, and an NPA of 0.93 and 0.85 on the QuantStudio™ 5 and BaseTyper™ 48.4, respectively. The corresponding 2x2 table is shown in **Table 3**. Thus, 17 and 19 samples were found to be methylated with the EpiDirect^®^ MGMT Assay when using the QuantStudio™ 5 and BaseTyper™ 48.4 instrument, respectively, vs. 18 samples when using the *therascreen*^*®*^ MGMT Kit. Similarly, 28 and 26 samples were found to be unmethylated with the EpiDirect^®^ MGMT Assay on the two different qPCR instruments vs. 27 with the *therascreen*^*®*^ MGMT Kit.

**Table 3.** 2×2 table comparing *MGMTp* methylation results obtained for Cohort 1 with the EpiDirect^®^ MGMT Assay and the *therascreen*^*®*^ MGMT Kit. A 5% methylation percentage cutoff and standard curve-derived algorithms were used with the EpiDirect^®^ MGMT Assay, and a 10% methylation percentage cutoff was used for the *therascreen*^*®*^ MGMT Kit.

| **Device** |  |  | ***therascreen*<sup>®</sup> MGMT Kit** |  |  |
|---|---|---|---|---|---|
|  |  |  | Methylated | Unmethylated | Total |
| **EpiDirect<sup>®</sup> MGMT Assay** | **QuantStudio™ 5** | Methylated | 15 | 2 | 17 |
|  |  | Unmethylated | 3 | 25 | 28 |
|  |  | Total | **18** | **27** | **45** |
|  | **BaseTyper™ 48.4** | Methylated | 15 | 4 | 19 |
|  |  | Unmethylated | 3 | 23 | 26 |
|  |  | Total | **18** | **27** | **45** |

This 5% cutoff is compared visually to the cutoff used with the *therascreen*^*®*^ MGMT Kit in **Figure 5**. It can be appreciated from the figure that several of the discrepant cases are close to either or both cutoffs used. Notably, the three samples that are methylated with the *therascreen*^*®*^ MGMT Kit cutoff but unmethylated with EpiDirect^®^ MGMT Assay all show a relatively high variation in pyrosequencing between the different CpG sites and, in particular CpG 79, which is not included in the EpiDirect^®^ MGMT Assay, shows an above-average methylation percentage (**Supplementary Table 5**). Furthermore, one of the cases found methylated with the EpiDirect^®^ MGMT Assay but unmethylated with the *therascreen*^*®*^ MGMT Kit (Sample 20) had two individual CpG sites with pyrosequencing methylation percentages at or above the applied average 10% cutoff (10% and 12%, respectively).

**Figure 5.**
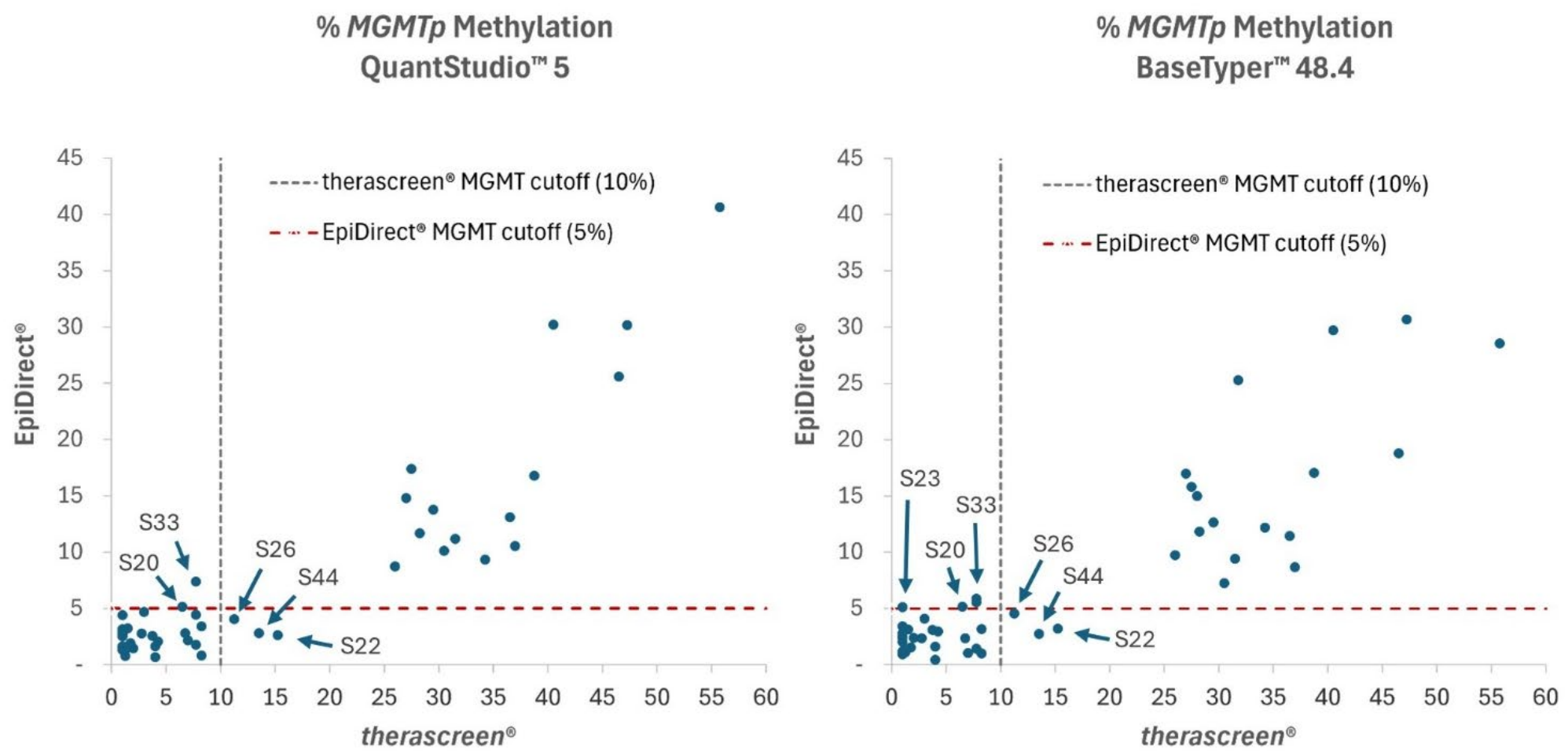
Correlation between determined methylation percentages of the *MGMT* promoter (*MGMTp*) by the EpiDirect^®^ MGMT Methylation qPCR Assay and the *therascreen*^®^ MGMT Kit in Cohort 1, including cutoffs as indicated. The EpiDirect^®^ MGMT Methylation qPCR Assay was used with the QuantStudio™ 5 Real-Time PCR System (left) and the BaseTyper™ 48.4 Real-Time PCR System (right). *MGMTp* methylation was calculated using the algorithm generated from including a standard curve in each run. Discrepant sample numbers are shown.

#### Impact of decreasing DNA input

The impact of decreasing DNA input on the determined *MGMTp* methylation percentages by the EpiDirect^®^ MGMT Assay was investigated by diluting selected samples in nuclease-free water. As shown in **Figure 6**, the calculated *MGMTp* methylation percentages did not significantly change when diluting three different patient samples from 25 ng per reaction to 2.5 ng per reaction.

**Figure 6.**
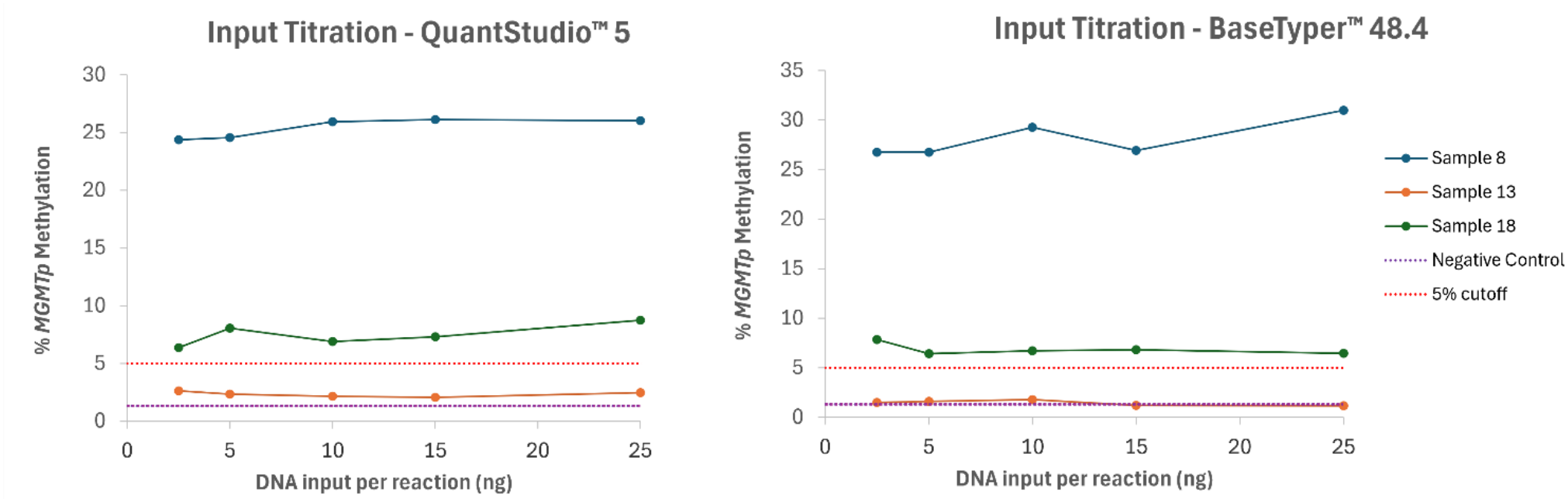
Titration of DNA input using Cohort 1 samples 8, 13 and 18. Calculated *MGMT* promoter (*MGMTp*) methylation percentages as determined by the EpiDirect^®^ MGMT Assay using the QuantStudio™ 5 (left) and the BaseTyper™ 48.4 (right) Real-Time PCR System are depicted as a function of the amount of DNA input as measured by the Qubit Fluorometer (Qiagen). *Average methylation percentage of two qPCR runs. Within sample P value > 0.21 using 1-way ANOVA.

### Cohort 2

#### Analysis of purified DNA from fresh frozen samples

To further investigate the correlation between EpiDirect^®^ MGMT Assay and *therascreen*^*®*^ MGMT Kit results in the clinical setting, we retrospectively evaluated 33 fresh frozen brain tumor samples where the methylation status had previously been determined using the *therascreen*^*®*^ MGMT Kit also with a 10% average *MGMTp* methylation cutoff (**Supplementary Table 2**). In this study, no standard curve controls were included, and the standard IFU algorithm was used for calculating corresponding methylation percentages.

Similar to cohort 1, the EpiDirect^®^ MGMT Assay results correlated well with the *therascreen*^*®*^ MGMT Kit results in cohort 2 with a slope of 0.60 and an R^2^ value of 0.83 (**Figure 7**). A ROC curve analysis with Youden’s Index found an optimal cutoff for the EpiDirect^®^ MGMT Assay to be 4.8% methylation when using a 10% methylation cutoff for the *therascreen*^*®*^ MGMT Kit (**Table 4**). This led to a PPA and an NPA of 1.00 and 0.95, respectively, and an overall percent agreement (OPA) of 0.97 (32 out of 33 cases). Thus, the optimal cutoff was also equal to that for obtaining similar PPA and NPA. The clinical site decided to use a methylation cutoff of 7% with the EpiDirect^®^ MGMT Assay in order to obtain 100% unmethylated sample agreement with the *therascreen*^*®*^ MGMT Kit, although this led to a lower overall percent agreement (**Figure 7** and **Table 4**). Thus, sample 7 was called non-methylated with this cutoff for the EpiDirect^®^ MGMT Assay (5.5% methylation vs 19% methylation with the *therascreen*^*®*^ MGMT Kit). Also, samples 10 and 32 that were found to be 12% and 13% methylated with the *therascreen*^*®*^ MGMT Kit, respectively, were called non-methylated with the EpiDirect^®^ MGMT Assay. These samples are, however, considered borderline methylated with the *therascreen*^*®*^ MGMT Kit and therefore do not support lowering the threshold for the EpiDirect^®^ MGMT Assay. Furthermore, both sample 7 and sample 32 had a markedly higher methylated CpG 79 compared to the remaining three included CpG sites (**Supplementary Table 5**). Consequently, when excluding CpG 79 (which is not targeted by the EpiDirect^®^ MGMT Assay), the mean methylation percentages for sample 7 and 32 were 10% and 5%, respectively, and thus much more in line with the EpiDirect^®^ MGMT Assay results. Also, sample 10 had one CpG site (CpG 77) with a markedly higher methylation percentage (34%) compared to the remaining three targeted CpG sites (7%, 4% and 7%) as determined by the *therascreen*^*®*^ MGMT Kit.

**Table 4.** Cohort 2 receiver operating characteristic (ROC) curve analysis for determination of corresponding cutoff when comparing EpiDirect^®^ MGMT Assay results on the QuantStudio™ Real-Time PCR System with the *therascreen*^®^ MGMT Kit. A cutoff of 10% methylation was used for the *therascreen®* MGMT Kit. *Based on ROC curve analysis with Youden’s index. The optimal cutoff was also identical to the cutoff needed for obtaining similar PPA and NPA. PPA, positive percent agreement (methylated samples). NPA, negative percent agreement (unmethylated samples). OPA, overall percent agreement. AUC, Area Under the Curve.

| **QuantStudio™ 5** |  |  |  |  |  |
|---|---|---|---|---|---|
| **Method** | **Cutoff (% methylation)** | **PPA** | **NPA** | **OPA** | **AUC** |
| Optimal cutoff* | 4.8 | 1.00 (11/11) | 0.95 (21/22) | 0.97 (32/33) | 0.996 |
| Laboratory procedure | 7.0 | 0.72 (8/11) | 1.00 (22/22) | 0.91 (30/33) |  |

**Figure 7.**
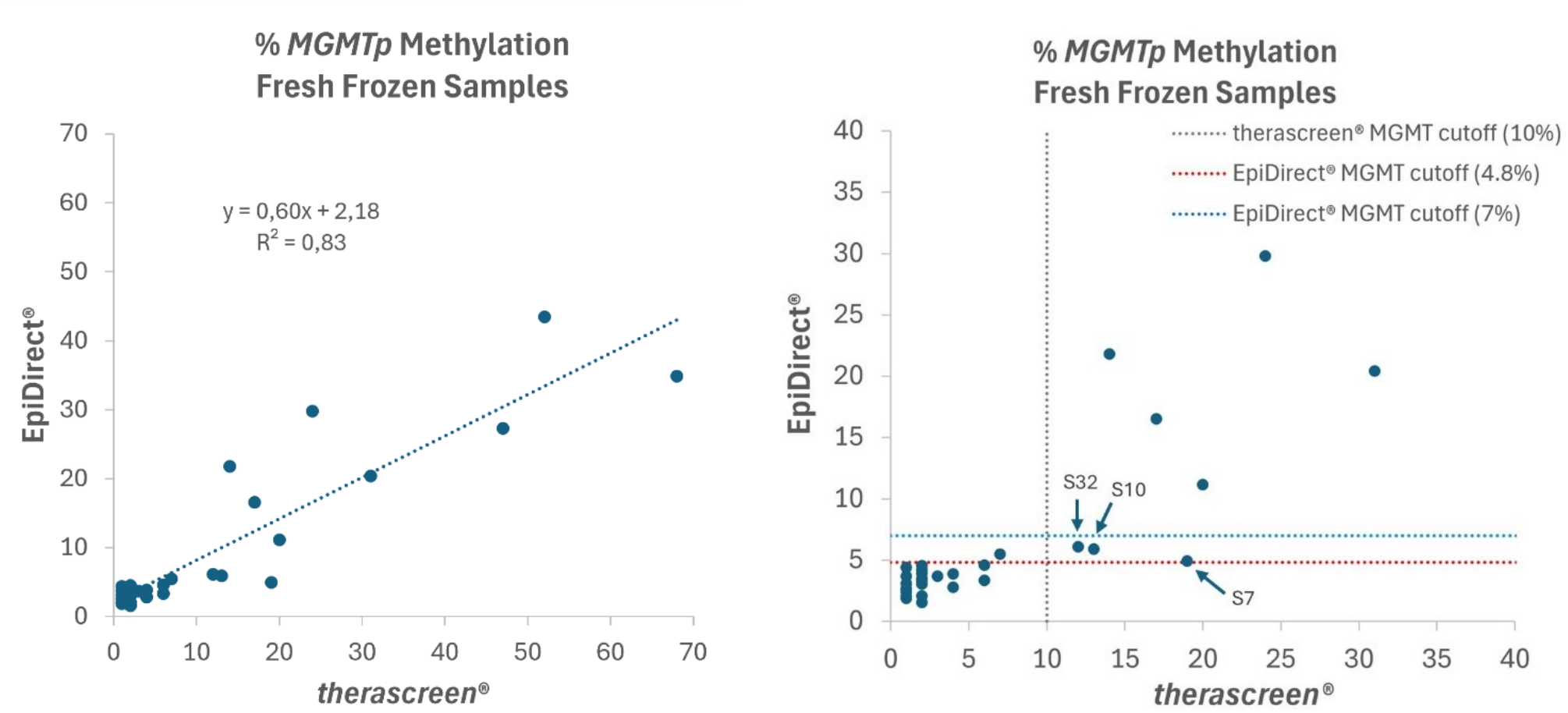
Correlation between determined methylation percentages of the *MGMT* promoter (*MGMTp*) by the EpiDirect^®^ MGMT Assay using the QuantStudio™ 5 Real-Time PCR System and the *therascreen*^®^ MGMT Kit when using purified DNA from fresh frozen samples from Cohort 2. **Left**: Resulting trendline and linear regression analysis. **Right**: Illustration of applied cutoffs and discrepant cases excluding samples with methylation percentages above 40%.

The corresponding 2×2 table when using a 7% methylation cutoff with the EpiDirect^®^ MGMT Assay is shown in **Table 5**.

**Table 5.**
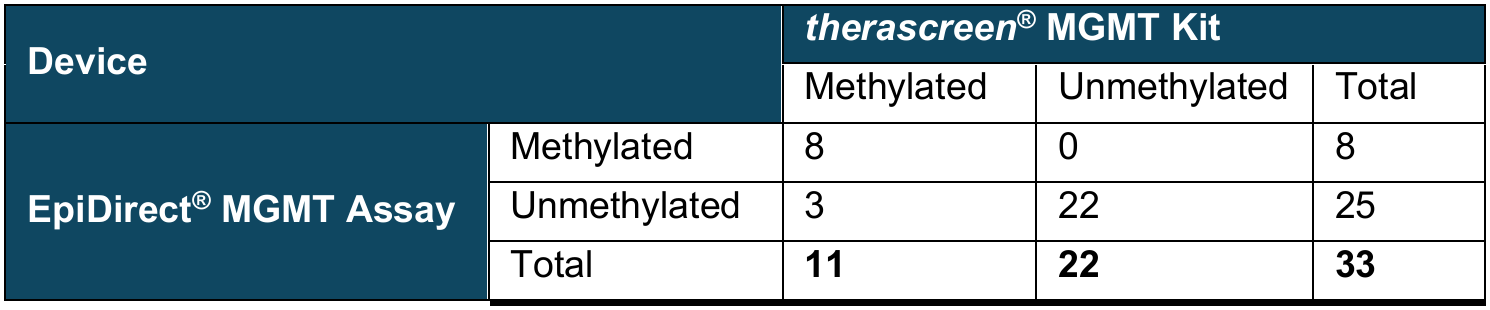
2×2 table comparing Cohort 2 *MGMT* promoter methylation results obtained with the EpiDirect^®^ MGMT Assay and the *therascreen*^*®*^ MGMT Kit. A 7% methylation percentage cutoff calculated using the IFU algorithm was used with the EpiDirect^®^ MGMT Assay, and a 10% methylation percentage cutoff was used with the *therascreen*^®^ MGMT Kit.

| **Device** |  | ***therascreen*<sup>®</sup> MGMT Kit** |  |  |
|---|---|---|---|---|
|  |  | Methylated | Unmethylated | Total |
| **EpiDirect<sup>®</sup> MGMT Assay** | Methylated | 8 | 0 | 8 |
|  | Unmethylated | 3 | 22 | 25 |
|  | Total | **11** | **22** | **33** |

## Discussion

The principal aim of this study was to evaluate the degree of correlation between the EpiDirect^®^ MGMT Assay and the *therascreen*^*®*^ MGMT Kit and to define corresponding cutoff values for the former, using the established thresholds of the latter as a reference. A high degree of correlation was observed between the two methods, despite their fundamentally different approaches to quantifying *MGMTp* methylation. Notably, the determined *MGMTp* methylation percentages and corresponding cutoffs were generally lower with the EpiDirect^®^ MGMT Assay. This may be due to the pre-treatment with bisulfite required for pyrosequencing, which has been reported to result in methylation-dependent degradation of DNA and consequently overestimation of methylation percentage. Alternatively, it can, at least in part, be explained by loss of chromosome 10 heterozygosity, which is common in glioblastoma and on which the *MGMT* gene is found^33^. This will not affect pyrosequencing results but will impact the EpiDirect^®^ results because it uses the *TBP* gene on chromosome 6 as the target for the Internal Reference assay.

Technical cutoffs and clinical implications have previously been described for the *therascreen*^*®*^ MGMT Kit^34,35^. In this study, a 10% average methylation of the evaluated CpG sites was used with the *therascreen*^®^ MGMT Kit. While treatment data for the patients included in this study were unavailable and therapeutic decisions remained unaffected, the relatively high correlation between EpiDirect^®^ MGMT Assay and *therascreen*^*®*^ MGMT Kit results is expected to lead to similar treatment efficiencies when based on the identified corresponding EpiDirect^®^ MGMT Assay cutoffs. However, future studies should investigate the clinical significance of the applied cutoff values for the EpiDirect^®^ MGMT Assay in relation to treatment outcomes, with particular focus on cases exhibiting discordant methylation status between the EpiDirect^®^ MGMT Assay and comparator methods.

ROC curve analysis with Youden’s index was performed to identify corresponding cutoffs for the EpiDirect^®^ MGMT Assay. In cohort 1, the optimal cutoffs were found to be in the range of 5.9% to 8.2% methylation but also tended to prioritize NPA (methylation negative) over PPA (methylation positive) samples. When optimizing for equal NPA and PPA, the range was 3.1% to 6.1% methylation for the two different real-time PCR platforms when using the IFU algorithm for calculating sample methylation percentages and a much narrower range of 4.5% to 4.6% methylation when using standard curve algorithms. The clinical site decided to use a 5% cutoff with standard curve algorithms which led to an overall agreement with the *therascreen*^®^ MGMT Kit results of 84%. This is in line with a recent study by Németh *et al*. also using a 5% methylation cutoff for the EpiDirect^®^ MGMT Assay and finding an overall agreement with pyrosequencing of 88.6% in IDH-wildtype glioblastoma samples^36^.

In cohort 2, which was based on fresh frozen samples and a pre-defined algorithm for calculating *MGMT* methylation, the optimal cutoff for the EpiDirect^®^ MGMT Assay (4.8% methylation) was also identical to the one for obtaining equal NPA and PPA values. The clinical site decided to use a cutoff of 7% in order to obtain 100% NPA which led to an overall agreement with therascreen^®^ MGMT Kit results of 91%.

The relatively few patients where the two methods did not reach the same result (n=9 in the two cohorts combined) can in most cases be associated with differences in targeted CpG sites or to being borderline cases. Thus, two of the nine discrepant cases had a much higher methylated CpG 79, which is not targeted by the EpiDirect^®^ MGMT Assay, compared to the remaining three CpG sites as determined by the *therascreen*^*®*^ MGMT Kit. Furthermore, five of the nine cases can be considered borderline since three were 7.8%, 11.3%, and 13% methylated, respectively, with the *therascreen*^*®*^ MGMT Kit (10% cutoff), and two samples in Cohort 1 were 5.1% and 5.2% methylated, respectively, with the EpiDirect^®^ MGMT Assay (5% cutoff). The remaining two samples were clearly unmethylated with the EpiDirect^®^ MGMT Assay while being above but still relatively close to the methylation percentage threshold with the *therascreen*^®^ MGMT Kit (13.5% and 15.3% methylated, respectively).

The second aim of this study was to investigate the correlation of EpiDirect^®^ MGMT Assay performance across real-time PCR systems and the impact of including a standard curve. In general, the correlation between data obtained with the QuantStudio™ 5 and BaseTyper™ 48.4 real-time PCR systems was very high. Using a standard curve for the quantitative analysis did not improve correlation between the two real-time PCR instruments when including all samples above the limit of detection but did improve correlation when only looking at samples with methylation percentages below 20%. The resulting slope of 0.97 shows that by using a standard curve, EpiDirect^®^ MGMT Assay data are essentially identical between the different real-time systems when evaluating samples with methylation percentages in the area of clinical relevance.

In summary, this study validates the EpiDirect^®^ MGMT Methylation qPCR Assay, a rapid method for measuring *MGMTp* methylation directly from untreated DNA, against the established *therascreen*^®^ MGMT Pyro Kit in brain tumor samples. Across FFPE and fresh frozen cohorts, the EpiDirect^®^ MGMT Assay showed solid agreement with pyrosequencing (R^2^ = 0.78-0.83), although it consistently reported lower methylation percentages. The results suggest that EpiDirect^®^ is a reliable and faster alternative to bisulfite-based *MGMTp* methylation testing for brain cancer diagnostics.

## Data Availability

All data produced in the present work are contained in the manuscript

## Acknowledgements

We would like to acknowledge Katrine Vogt Møller, Maja Isabella Hunt, and Jesper Bertelsen from the Department of Pathology, Aarhus University Hospital, Aarhus, Denmark, and Zaineb Adel Kamel Ali Al-Jaberi and Maiken Sønder Zacho from the Department of Pathology, Rigshospitalet, Copenhagen, Denmark, for their valuable technical assistance and contributions to the validation work.

## Footnotes

a In some cohort 2 runs, the positive control was replaced by a 5% control which was used for setting the threshold.

## References

1. Masood, S., Shaikh, A., and Brar, Y. (2025). Evaluating Molecular and Immune Biomarkers in Predicting Bladder Cancer Recurrence and Survival: A Systematic Review. Cureus. 10.7759/cureus.96824.

2. Xie, J., and Soleimani Samarkhazan, H. (2025). Beyond the DNA sequence: mapping the dynamic epigenetic landscape for risk stratification and therapeutic intervention in acute myeloid leukemia. Preprint, https://doi.org/10.1007/s10238-025-01972-w z10.1007/s10238-025-01972-w.

3. Ma, C., Cheng, J., Gu, J., and Wang, Q. (2025). Epigenetic drugs in cancer therapy: mechanisms, immune modulation, and therapeutic applications. Molecular Biomedicine 6, 132. 10.1186/s43556-025-00373-5.

4. Belanich, M., Pastor. M, Randall, T., Guerra, D., Kibitel, J., Alas, L., Li, B., Citron, M., Wasserman, P., White, A., et al. (1996). Retrospective study of the correlation between the DNA repair protein alkyltransferease and survival of brain tumor patients treated with carmustine. Cancer Res. 56, 783–788.

5. Jaeckle, K.A., Eyre, H.J., Towsend, J.J., Schulman, S., Knudson, H.M., Belanich, M., Yarosh, D.B., Bearman, S.I., Giroux, D.J., and Schold, S.C. (1998). Correlation of tumor O6 methylguanine-DNA methyltransferase levels with survival of malignant astrocytoma patients treated with bis-chloroethylnitrosurea: a Southwest Oncology Group study. Journal of Clinical Oncology 16, 3310–3315.

6. Friedman, H.S., McLendon, R.E., Kerby, T., Dugan, M., Bigner, S.H., Henry, A.J., Ashley, D.M., Krischer, J., Lowell, S., Rasheed, K., et al. (1998). DNA mismatch repair and O6-alkylguanine-DNA alkyltransferase analysis and response to Temodal in newly diagnosed malignant glioma. Journal of Clinical Oncology 16, 3851–3857.

7. Hegi, M.E., Diserens, A.-C., Godard, S., Dietrich, P.-Y., Regli, L., Ostermann, S., Otten, P., Melle, G. Van, Tribolet, N. de, and Stupp, R. (2004). Clinical Trial Substanciates the Predictive Value of O-6-Methylguanine-DNA Methyltransferase Prmooter Methylation in Glioblastoma PAtients Treated with Temozolomide. Clinical Cancer Research 10, 1871–1874.

8. Esteller, M., Garcia-Foncillas, J., Andion, E., Goodman, S.N., Hidalgo, O.F., Vanaclocha, V., Baylin, S.B., and Herman, J.G. (2000). Inactivation of the DNA-Repair Gene MGMT and the Clinical Response of Gliomas to Alkylating Agents. N. Engl. J. Med. 343, 1350–1354.

9. Stupp, R., Mason, W.P., van den Bent, M.J., Weller, M., Fischer, B., Taphoorn, M.J.B., Belanger, K., Brandes, A.A., Marosi, C., Bogdahn, U., et al. (2005). Radiotherapy plus concomitant and adjuvant temozolomide for glioblastoma. N. Engl. J. Med. 325, 987–996.

10. Gilbert, M.R., Wang, M., Aldape, K.D., Stupp, R., Hegi, M.E., Jaeckle, K.A., Armstrong, T.S., Wefel, J.S., Won, M., Blumenthal, D.T., et al. (2013). Dose-dense temozolomide for newly diagnosed glioblastoma: A randomized phase III clinical trial. Journal of Clinical Oncology 31, 4085–4091. 10.1200/JCO.2013.49.6968.

11. Paz, M.F., Yaya-Tur, R., Rojas-Marcos, I., Reynes, G., Pollan, M., Aguirre-Cruz, L., García-Lopez, J.L., Piquer, J., Safont, M.J., Balaña, C., et al. (2004). CpG island hypermethylation of the DNA repair enzyme methyltransferase predicts response to temozolomide in primary gliomas. Clin. Cancer Res. 10, 4933–4938. 10.1158/1078-0432.CCR-04-0392.

12. Hegi, M.E., Diserens, A.-C., Gorlia, T., Hamou, M.-F., Tribolet, N. de, Weller, M., Kros, J.M., Hainfeller, J.A., Mason, W., Mariani, L., et al. (2005). MGMT Gene Silencing and Benefit from Temozolomide in Glioblastoma. N. Engl. J. Med. 352, 997–1003.

13. Perry, J.R., Laperriere, N., O’Callaghan, C.J., Brandes, A.A., Menten, J., Phillips, C., Fay, M., Nishikawa, R., Cairncross, J.G., Roa, W., et al. (2017). Short-Course Radiation plus Temozolomide in Elderly Patients with Glioblastoma. N. Engl. J. Med. 376, 1027–1037. 10.1056/NEJMOA1611977.

14. Dunn, J., Baborie, A., Alam, F., Joyce, K., Moxham, M., Sibson, R., Crooks, D., Husband, D., Shenoy, A., Brodbelt, A., et al. (2009). Extent of MGMT promoter methylation correlates with outcome in glioblastomas given temozolomide and radiotherapy. Br. J. Cancer 101, 124–131. 10.1038/SJ.BJC.6605127.

15. Siller, S., Lauseker, M., Karschnia, P., Niyazi, M., Eigenbrod, S., Giese, A., and Tonn, J.C. (2021). The number of methylated CpG sites within the MGMT promoter region linearly correlates with outcome in glioblastoma receiving alkylating agents. Acta Neuropathol. Commun. 9, 35. 10.1186/S40478-021-01134-5.

16. Stupp, R., Brada, M., van den Bent, M.J., Tonn, J.C., and Pentheroudakis, G. (2014). High-grade glioma: ESMO clinical practice guidelines for diagnosis, treatment and follow-up. Annals of Oncology 25, 93–101. 10.1093/annonc/mdu050.

17. Burt Nabors, L., Portnow, J., Bhatia, A., Bloch, O., Brem, S., Butowski, N., Cannon, D.M., Chao, S.T., Chheda, M.G., Fabiano, A.J., et al. (2025). NCCN Clinical Practice Guidelines in Oncology (NCCN Guidelines®) -Central Nervous System Cancers Version 3. 2025.

18. Bienkowski, M., Berghoff, A.S., Marosi, C., Wöhrer, A., Heinzl, H., Hainfellner, J.A., and Preusser, M. (2015). Clinical Neuropathology practice guide 5-2015: MGMT methylation pyrosequencing in glioblastoma: unresolved issues and open questions. Clin. Neuropathol. 34, 250–257. 10.5414/NP300904.

19. Malley, D.S., Hamoudi, R.A., Kocialkowski, S., Pearson, D.M., Collins, V.P., and Ichimura, K. (2011). A distinct region of the MGMT CpG island critical for transcriptional regulation is preferentially methylated in glioblastoma cells and xenografts. Acta Neuropathol. 121, 651–661.

20. Nakagawachi, T., Soejima, H., Urano, T., Zhao, W., Higashimoto, K., Satoh, Y., Matsukura, S., Kudo, S., Kitajima, Y., Harada, H., et al. (2003). Silencing e?ect of CpG island hypermethylation and histone modifications on O6-methylguanine-DNA methyltransferase (MGMT) gene expression in human cancer. Oncogene 22, 8835–8844. 10.1038/SJ.ONC.1207183.

21. Shah, N., Lin, B., Sibenaller, Z., Ryken, T., Lee, H., Yoon, J.G., Rostad, S., and Foltz, G. (2011). Comprehensive analysis of MGMT promoter methylation: correlation with MGMT expression and clinical response in GBM. PLoS One 6. 10.1371/JOURNAL.PONE.0016146.

22. Leske, H., Camenisch Gross, U., Hofer, S., Neidert, M.C., Leske, S., Weller, M., Lehnick, D., and Rushing, E.J. (2023). MGMT methylation pattern of long-term and short-term survivors of glioblastoma reveals CpGs of the enhancer region to be of high prognostic value. Acta Neuropathol. Commun. 11. 10.1186/S40478-023-01622-W.

23. Herman, J.G., Gră, J.R., Myöhänen, S., Nelkin, B.D., and Baylin, S.B. (1996). Methylation-specific PCR: a novel PCR assay for methylation status of CpG islands. Proc. Natl. Acad. Sci. U. S. A. 93, 9821–9826. 10.1073/PNAS.93.18.9821.

24. Colella, S., Shen, L., Baggerly, K.A., Issa, J.P.J., and Krahe, R. (2003). Sensitive and quantitative universal Pyrosequencing methylation analysis of CpG sites. Biotechniques 35, 146–150. 10.2144/03351MD01.

25. Nygren, A.O.H., Ameziane, N., Duarte, H.M.B., Vijzelaar, R.N.C.P., Waisfisz, Q., Hess, C.J., Schouten, J.P., and Errami, A. (2005). Methylation-Specific MLPA (MS-MLPA): Simultaneous detection of CpG methylation and copy number changes of up to 40 sequences. Nucleic Acids Res. 33, 1–9. 10.1093/nar/gni127.

26. Halldorsson, S., Nagymihaly, R.M., Patel, A., Brandal, P., Panagopoulos, I., Leske, H., Lund-Iversen, M., Sahm, F., and Vik-Mo, E.O. (2024). Accurate and comprehensive evaluation of O6-methylguanine-DNA methyltransferase promoter methylation by nanopore sequencing. Neuropathol. Appl. Neurobiol. 50. 10.1111/nan.12984.

27. Olova, N., Krueger, F., Andrews, S., Oxley, D., Berrens, R. V., Branco, M.R., and Reik, W. (2018). Comparison of whole-genome bisulfite sequencing library preparation strategies identifies sources of biases a?ecting DNA methylation data. Genome Biol. 19. 10.1186/S13059-018-1408-2.

28. Bendixen, K.K., Mindegaard, M., Epistolio, S., Dazio, G., Marchi, F., Spina, P., Arnspang, E.C., Soerensen, M., Christensen, U.B., Frattini, M., et al. (2023). A qPCR technology for direct quantification of methylation in untreated DNA. Nat. Commun. 14. 10.1038/S41467-023-40873-Y.

29. Christensen, U.B., and Pedersen, E.B. (2002). Intercalating nucleic acids containing insertions of 1-O -(1-pyrenylmethyl)glycerol: stabilisation of dsDNA and discrimination of DNA over RNA. Nucleic Acids Res. 30, 4918–4925. 10.1093/NAR/GKF624.

30. Christensen, U.B., Wamberg, M., El-Essawy, F.A.G., Ismail, A.E.H., Nielsen, C.B., Filichev, V. V., Jessen, C.H., Petersen, M., and Pedersen, E.B. (2004). Intercalating Nucleic Acids: The Influence of Linker Length and Intercalator Type on Their Duplex Stabilities. Nucleosides Nucleotides Nucleic Acids 23, 207–225. 10.1081/NCN-120027829.

31. Nielsen, C.B., Petersen, M., Pedersen, E.B., Hansen, P.E., and Christensen, U.B. (2004). NMR Structure Determination of a Modified DNA Oligonucleotide Containing a New Intercalating Nucleic Acid. Bioconjug. Chem. 15, 260–269.

32. Christensen, U.B., and Pedersen, E.B. (2003). Intercalating Nucleic Acids with pyrene nucleotide analogues as next-nearest neihbours for excimer fluorescence detection of single point mutations under nonstringent hybridisation conditions. Helv. Chim. Acta 86, 2090–2097.

33. Richard, S., Tachon, G., Milin, S., Wager, M., and Karayan-Tapon, L. (2020). Dual MGMT inactivation by promoter hypermethylation and loss of the long arm of chromosome 10 in glioblastoma. Cancer Med. 9, 6344–6353. 10.1002/cam4.3217.

34. Skarin, N., Hallbeck, M., Werlenius, K., Rohan, Z., Sandström, M., Turanyi, E., Löfgren, D., Johansson, H., Strandéus, M., Tavelin, B., et al. (2026). MGMT promoter methylation status for glioblastoma: defining the clinically relevant cut-offvalue for pyrosequencing. J. Neurooncol. 178. 10.1007/s11060-026-05633-0.

35. Quillien, V., Lavenu, A., Ducray, F., Meyronet, D., Chinot, O., Fina, F., Sanson, M., Carpentier, C., Karayan-Tapon, L., Rivet, P., et al. (2017). Clinical validation of the CE-IVD marked Therascreen MGMT kit in a cohort of glioblastoma patients. Cancer Biomarkers 20, 435–441. 10.3233/CBM-170191.

36. Németh, A., Selignow, C., May, A., Schmid, K., Dohmen, H., Stein, M., Trautner, J., Haag, D., Sahm, F., and Acker, T. (2026). EpiDirect enables rapid bisulfite-free MGMT promoter methylation testing in IDH-wildtype glioblastoma with high agreement across established assays. Neurooncol. Adv. 8, vdag255. 10.1093/NOAJNL/VDAG255.

