## Supplementary information for "Quantitative evaluation of *MGMT* promoter methylation in FFPE and fresh frozen brain tumor samples by EpiDirect MGMT Methylation qPCR Assay compared to pyrosequencing: correlation and corresponding cutoffs"

| Sample | Gender | Age range | %TC | Diagnosis | Sample | Gender | Age range | %TC | Diagnosis |
|---|---|---|---|---|---|---|---|---|---|
| 1 | M | 70-79 | 50 | G | 25 | M | 60-69 | 80 | G |
| 2 | F | 60-69 | 70 | G | 26 | M | 60-69 | 80 | G |
| 3 | M | 50-59 | 60 | G | 27 | M | 40-49 | 80 | G |
| 4 | F | 60-69 | 70 | G | 28 | M | 50-59 | 60 | G |
| 5 | F | 60-69 | 70 | G | 29 | M | 40-49 | 60 | O |
| 6 | F | 70-79 | 60-70 | G | 30 | F | 50-59 | 80 | G |
| 7 | F | 60-69 | 60 | O | 31 | M | 70-79 | 80 | G |
| 8 | M | 50-59 | 60-70 | G | 32 | M | 50-59 | 40 | G |
| 9 | F | 60-69 | 70-80 | $O^1$ | 33 | M | 70-79 | 40 | G |
| 10 | M | 60-69 | 50-60 | G | 34 | F | 70-79 | 80 | G |
| 11 | M | 50-59 | 50-60 | G | 35 | F | 50-59 | 80 | G |
| 12 | F | 60-69 | 70 | G | 36 | M | 50-59 | 70 | G |
| 13 | F | 70-79 | 80 | G | 37 | M | 70-79 | 80 | G |
| 14 | M | 60-69 | 70 | G | 38 | M | 70-79 | 80 | G |
| 15 | M | 40-49 | 80 | $G^1$ | 39 | M | 50-59 | 75 | G |
| 16 | F | 60-69 | 80 | G | 40 | M | 20-29 | 70 | D |
| 17 | F | 70-79 | 20 | C | 41 | F | 60-69 | 60 | $C^1$ |
| 18 | F | 60-69 | 80 | G | 42 | F | 40-49 | 80 | G |
| 19 | M | 60-69 | 70 | $O^1$ | 43 | M | 70-79 | 50 | G |
| 20 | F | 60-69 | 50-60 | G | 44 | F | 80-89 | 60 | G |
| 21 | M | 50-59 | 80 | $G^2$ | 45 | M | 70-79 | 80 | G |
| 22 | F | 60-69 | 70 | G | 46* | M | NA | 50 | O |
| 23 | M | 30-39 | 60-70 | G | 47* | M | NA | 50 | O |
| 24 | F | 60-69 | 80 | G | | | | | |

**Supplementary Table 1.** Cohort 1 clinical-pathological data for included brain cancer patients. All samples are from cerebrum. %TC: Tumor cell percentage. G) Glioblastoma, IDH wildtype, WHO grade 4, $G^1$) with sarcomatoid component, $G^2$) with primitive neuronal component, O) Oligodendroglioma with IDH-mutation and 1p/19q co-deletion, WHO grade 2, $O^1$) WHO grade 3, C) Glioma with high malignancy, $C^1$ adult-type diffuse glioma, WHO grade 4, IDH wildtype, subtype F, D) Unspecific alteration. *External Quality Assessment (EQA) sample from GenQA.

| Sample | Gender | Age range | Diagnosis | Sample | Gender | Age range | Diagnosis |
|---|---|---|---|---|---|---|---|
| 1 | M | 70-79 | G | 18 | M | 30-39 | A |
| 2 | F | 10-19 | A | 19 | F | 80-89 | G |
| 3 | M | 70-79 | G | 20 | M | 50-59 | G |
| 4 | F | 50-59 | G | 21 | F | 70-79 | G |
| 5 | M | 70-79 | G | 22 | M | 60-69 | G |
| 6 | M | 80-89 | O | 23 | M | 60-69 | G |
| 7 | M | 70-79 | G | 24 | M | 60-69 | G |
| 8 | F | 50-59 | G | 25 | M | 60-69 | G |
| 9 | F | 40-49 | G | 26 | M | 60-69 | G |
| 10 | F | 70-79 | G | 27 | M | 50-59 | G |
| 11 | F | 30-39 | A | 28 | M | 60-69 | G |
| 12 | M | 70-79 | G | 29 | F | 70-79 | G |
| 13 | M | 70-79 | G | 30 | M | 70-79 | G |
| 14 | M | 60-69 | G | 31 | M | 70-79 | G |
| 15 | F | 60-69 | G | 32 | F | 50-59 | A |
| 16 | F | 60-69 | G | 33 | F | 40-49 | G |
| 17 | F | 40-49 | A | | | | |

**Supplementary Table 2**. Cohort 2 clinical-pathological data for included brain cancer patients. All samples are from cerebrum. G) Glioblastoma, IDH wildtype, WHO grade 4, O) Oligodendroglioma with IDH-mutation and 1p/19q co-deletion, A) Astrocytoma, WHO grade 2.

| Sample | % *MGMTp* Methylation | | | | | Sample | % *MGMTp* Methylation | | | | |
|---|---|---|---|---|---|---|---|---|---|---|---|
| | *therascreen*® MGMT Kit | EpiDirect® MGMT Assay | | | | | *therascreen*® MGMT Kit | EpiDirect® MGMT Assay | | | |
| | | QuantStudio™ 5 | | BaseTyper™ 48.4 | | | | QuantStudio™ 5 | | BaseTyper™ 48.4 | |
| | | IFU[1] | StdC.[2] | IFU | Std C. | | | IFU | StdC. | IFU | StdC. |
| 1 | 1.0 | 0.7 | 1.3 | 1.4 | 0.9 | 25 | 46.5 | 25.2 | 25.6 | 27.8 | 18.8 |
| 2 | 8.3 | 0.4 | 0.8 | 1.5 | 1.0 | 26 | 11.4 | 2.9 | 4.0 | 6.8 | 4.6 |
| 3 | 1.0 | 1.9 | 3.2 | 4.4 | 3.4 | 27 | 4.0 | 1.0 | 1.6 | 2.4 | 1.6 |
| 4 | 31.5 | 7.6 | 11.2 | 11.1 | 9.4 | 28 | 8.3 | 2.6 | 3.4 | 4.3 | 3.2 |
| 5 | 36.5 | 9.0 | 13.1 | 13.2 | 11.4 | 29 | 27.0 | 14.9 | 14.8 | 18.8 | 17.0 |
| 6 | 40.5 | 22.7 | 30.2 | 31.3 | 29.7 | 30 | 1.5 | 2.4 | 3.2 | 4.2 | 3.1 |
| 7 | 30.5 | 6.8 | 10.1 | 8.8 | 7.2 | 31 | 28.3 | 11.2 | 11.7 | 13.7 | 11.8 |
| 8 | 55.8 | 31.4 | 40.6 | 35.2 | 28.5 | 32 | 1.0 | 1.0 | 1.6 | 1.8 | 1.2 |
| 9 | 3.0 | 2.9 | 4.7 | 5.6 | 4.1 | 33 | 7.8 | 6.5 | 7.4 | 7.4 | 5.9 |
| 10 | 1.0 | 1.7 | 2.9 | 3.4 | 2.4 | 34 | 4.0 | 0.4 | 0.7 | 0.7 | 0.4 |
| 11 | 1.0 | 1.5 | 2.5 | 2.9 | 2.0 | 35 | 3.8 | 1.5 | 2.5 | 3.8 | 3.1 |
| 12 | 37.0 | 7.1 | 10.5 | 11.4 | 8.7 | 36 | 7.8 | 3.1 | 4.4 | 6.5 | 5.6 |
| 13 | 1.8 | 1.1 | 1.9 | 2.2 | 1.5 | 37 | 7.0 | 1.3 | 2.2 | 1.4 | 1.1 |
| 14 | 27.5 | 12.3 | 17.4 | 20.1 | 15.8 | 38 | 7.8 | 1.0 | 1.8 | 1.8 | 1.4 |
| 15 | 1.0 | 1.5 | 1.5 | 2.5 | 2.8 | 39 | 38.8 | 15.7 | 16.8 | 18.8 | 17.1 |
| 16 | 2.0 | 1.4 | 1.5 | 2.1 | 2.4 | 40 | 1.3 | 0.7 | 1.3 | 1.6 | 1.2 |
| 17 | 4.3 | 2.0 | 2.1 | 2.7 | 3.0 | 41 | 6.8 | 1.8 | 2.8 | 2.9 | 2.4 |
| 18 | 26.0 | 8.0 | 8.7 | 9.3 | 9.8 | 42 | 2.8 | 1.9 | 2.7 | 3.3 | 2.4 |
| 19 | 29.5 | 12.5 | 13.8 | 12.2 | 12.6 | 43 | 1.0 | 1.9 | 2.7 | 3.5 | 2.5 |
| 20 | 6.5 | 4.9 | 5.2 | 4.8 | 5.1 | 44 | 13.5 | 2.0 | 2.8 | 3.8 | 2.8 |
| 21 | 1.3 | 0.8 | 0.8 | 0.9 | 1.1 | 45 | 47.3 | 31.7 | 30.2 | 33.1 | 30.7 |
| 22 | 15.3 | 1.8 | 2.6 | 4.7 | 3.2 | 46[3] | 28.0 | 11.1 | 12.2 | 17.7 | 15.0 |
| 23 | 1,0 | 3,2 | 4,4 | 7.6 | 5,1 | 47[3] | 31.8 | 22.2 | 20.0 | 27.8 | 25.3 |
| 24 | 34.3 | 7.7 | 9.3 | 18.0 | 12.2 | | | | | | |

**Supplementary Table 3.** *MGMT* promoter (*MGMTp*) methylation percentages of 47 brain tumor samples as determined by the *therascreen*® MGMT Kit and EpiDirect® MGMT Assay. [1+2]EpiDirect® MGMT Assay data are for each qPCR instrument shown using the instrument-specific algorithm according to the Instructions for Use (IFU) version 2.8 as well as the algorithm generated by including a standard curve (StdC). [3]External Quality Assessment (EQA) sample from GenQA. Green wells indicate MGMT methylation positive samples as determined by the therascreen® MGMT Kit when using a cutoff of 10% methylation.

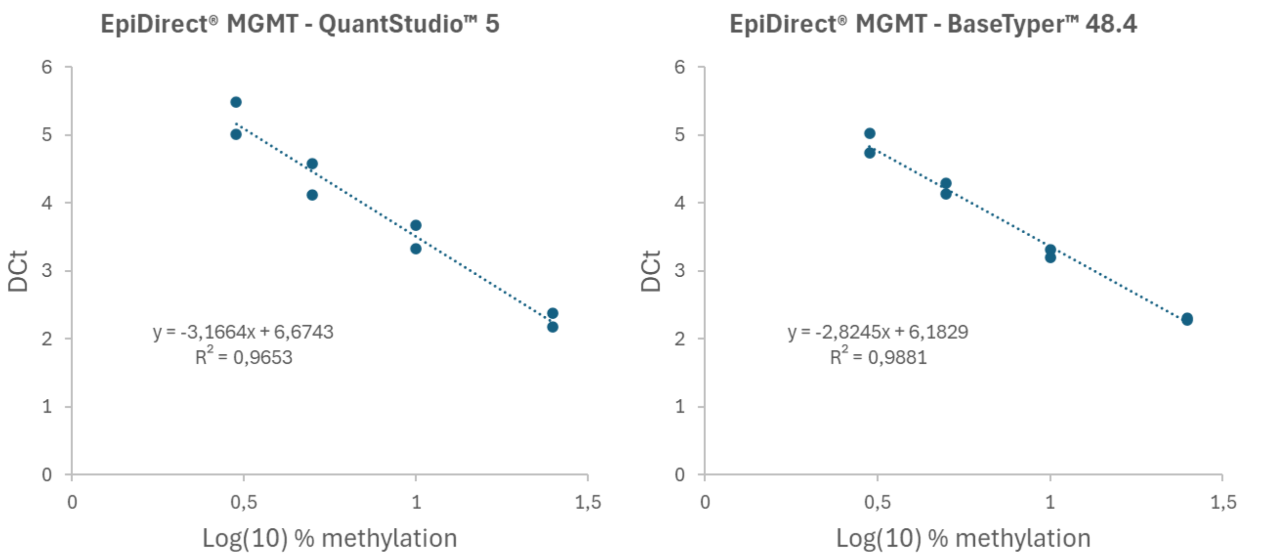


**Supplementary Figure 1**. Examples of standard curves and corresponding algorithms generated with the EpiDirect® MGMT Assay using the QuantStudio™ 5 (left) and BaseTyper™ 48.4 (right) real-time PCR systems. The standard curves were generated using duplicate reactions of the included positive control (25% methylation) as well as additional methylated human DNA controls (3%, 5% and 10% methylated human DNA).

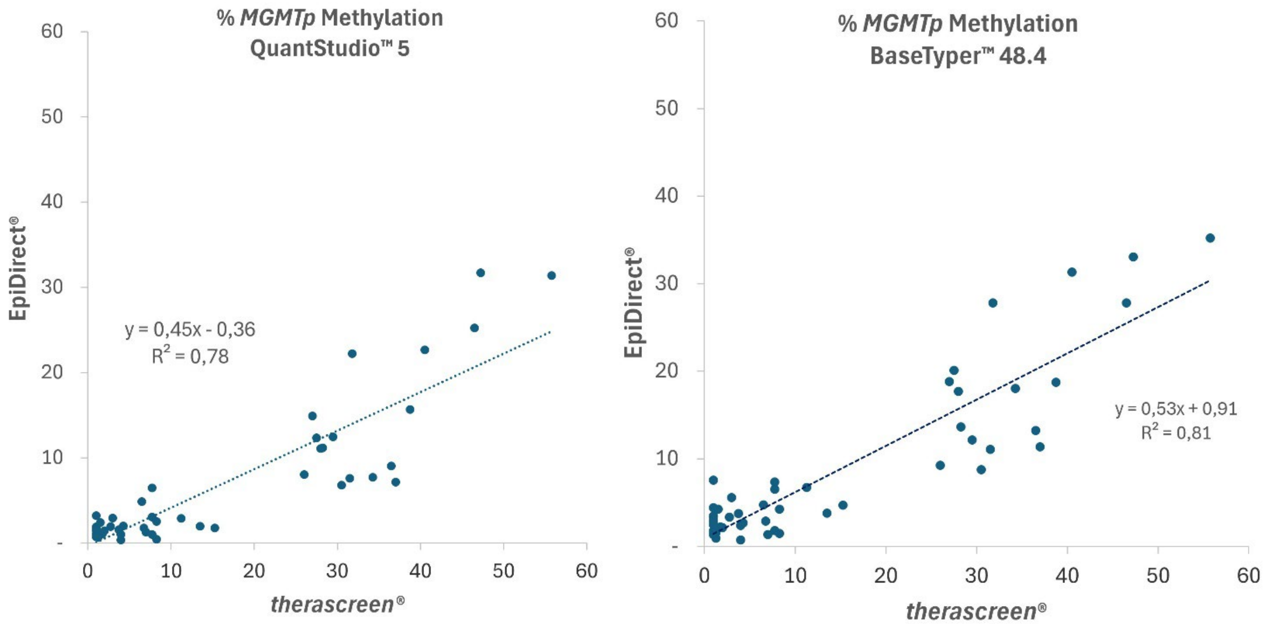


**Supplementary Figure 2.** Correlation between determined methylation percentages of the *MGMT* promoter (*MGMTp*) by the EpiDirect® MGMT Assay and the *therascreen*® MGMT Kit. The EpiDirect® MGMT Assay was used with the QuantStudio™ 5 (left) and the BaseTyper™ 48.4 Real-Time PCR System (right) and *MGMTp* methylation was calculated using the Instructions for Use (IFU) version 2.8 algorithms.

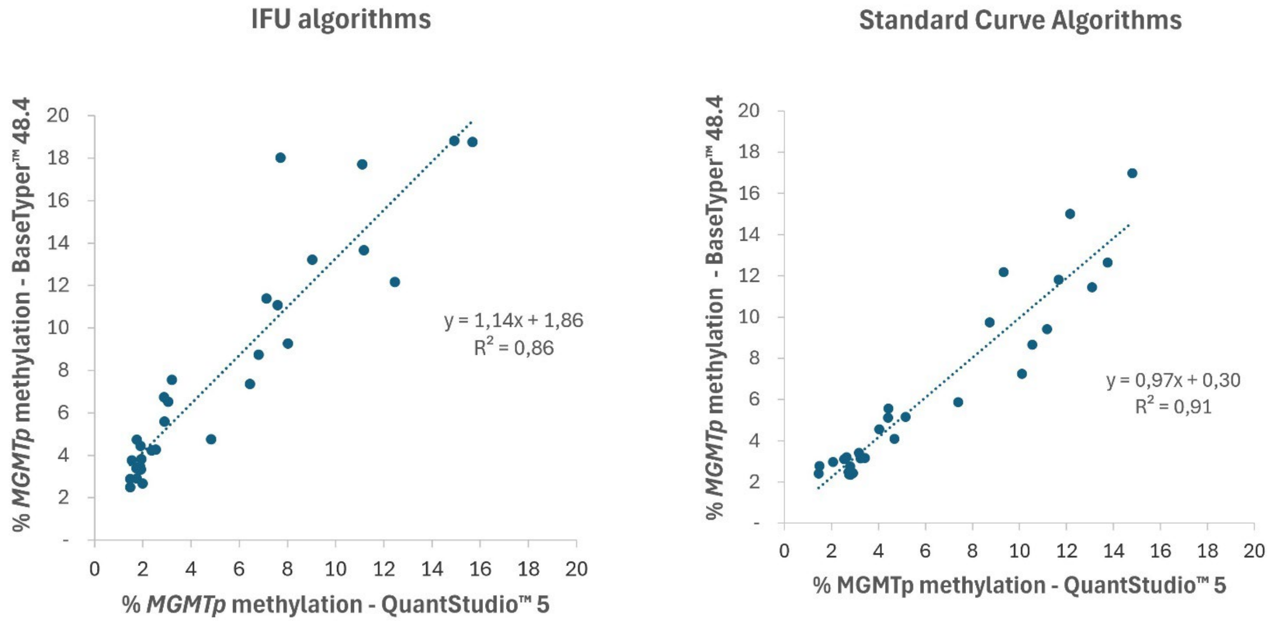


**Supplementary Figure 3.** Correlation between *MGMT* promoter (*MGMTp*) methylation percentages determined by the EpiDirect® MGMT Assay when using the QuantStudio™ 5 and the BaseTyper™ 48.4 Real-time PCR systems. Data are shown for *MGMTp* methylation percentages calculated using the algorithms described in the EpiDirect® MGMT Assay Instructions for Use (IFU) version 2.8 (left) and when including a standard curve in each run (right). Samples with *MGMTp* methylation above 20% and below the theoretical limit of detection on either real-time PCR system have been removed.

**Supplementary Figure 4**. ROC curve analysis with Youden's Index from comparison of EpiDirect® MGMT Assay results with the *therascreen*® MGMT Kit when using a 10% methylation cutoff. Data for the EpiDirect® MGMT Assay when using the QuantStudio™ 5 Real-Time PCR System is shown on the left and when using the BaseTyper™ 48.4 Real-Time PCR System on the right.

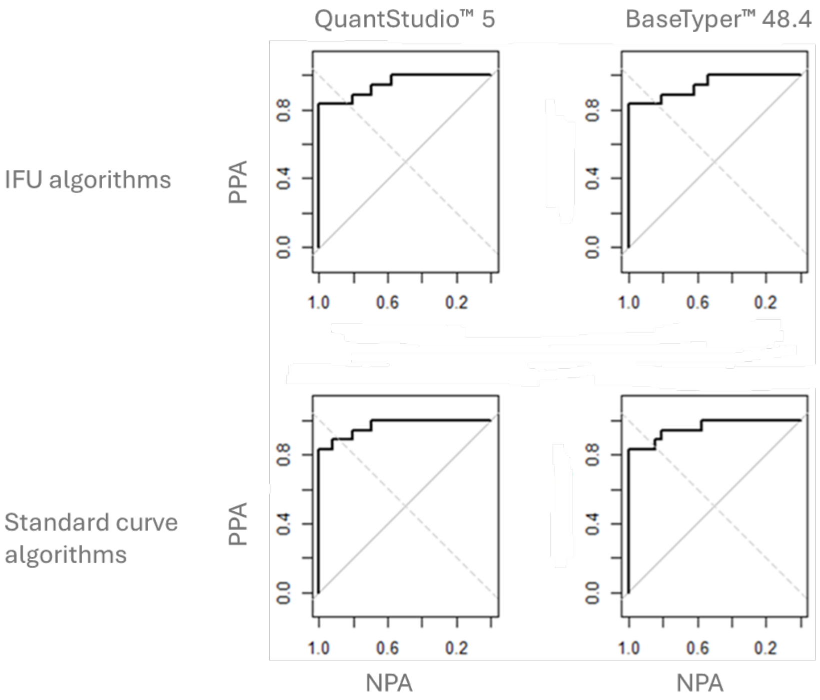


| Sample | % *MGMTp* Methylation EpiDirect® MGMT Assay | *therascreen*® MGMT Kit | Sample | EpiDirect® MGMT Assay | *therascreen*® MGMT Kit |
|---|---|---|---|---|---|
| 1 | 4.5 | 2 | 18 | 3.7 | 3 |
| 2 | 4.6 | 6 | 19 | 1.5 | 2 |
| 3 | 16.5 | 17 | 20 | 2.1 | 2 |
| 4 | 2.8 | 4 | 21 | 3.1 | 2 |
| 5 | 27.3 | 47 | 22 | 3.5 | 2 |
| 6 | 20.4 | 31 | 23 | 1.9 | 1 |
| 7 | 4.9 | 19 | 24 | 2.1 | 1 |
| 8 | 3.3 | 6 | 25 | 3.4 | 2 |
| 9 | 11.1 | 20 | 26 | 3.1 | 1 |
| 10 | 5.9 | 13 | 27 | 21.8 | 14 |
| 11 | 2.7 | 1 | 28 | 3.7 | 1 |
| 12 | 4.2 | 2 | 29 | 4.4 | 1 |
| 13 | 34.9 | 68 | 30 | 29.8 | 24 |
| 14 | 3.9 | 4 | 31 | 2.4 | 1 |
| 15 | 3.3 | 2 | 32 | 6.1 | 12 |
| 16 | 43.5 | 52 | 33 | 3.9 | 2 |
| 17 | 5.5 | 7 | | | |

**Supplementary Table 4.** *MGMT* promoter (*MGMTp*) methylation percentages of 33 fresh frozen brain tumor samples as determined by the *therascreen*® MGMT Kit and the EpiDirect® MGMT Assay using the QuantStudio™ 5 Real-Time PCR System. Green wells indicate MGMT methylation positive samples as determined by the therascreen® MGMT Kit when using a cutoff of 10% methylation.

| | % *MGMTp* Methylation | | | | | | |
|---|---|---|---|---|---|---|---|
| | *therascreen*® MGMT Kit | | | | | EpiDirect® MGMT Assay | |
| Sample | CpG 76 | CpG 77 | CpG 78 | CpG 79 | Average | QuantStudio™ 5 | BaseTyper™ 48.4 |
| **Cohort 1** | | | | | | | |
| *20* | 2 | 10 | 2 | 12 | **6.5** | **5.2** | **5.2** |
| *22* | 7 | 23 | 5 | 26 | **15.3** | **2.6** | **3.2** |
| *23* | 1 | 1 | 1 | 1 | **1** | **4.4 (not discrepant)** | **5.1** |
| *26* | 16 | 8 | 9 | 12 | **11.3** | **4.0** | **4.6** |
| *33* | 5 | 9 | 8 | 9 | **7.8** | **7.4** | **5.6** |
| *44* | 8 | 23 | 8 | 15 | **13.5** | **2.8** | **2.8** |
| **Cohort 2** | | | | | | | |
| *7* | 9 | 12 | 10 | 45 | **19.0** | **4.9** | **ND** |
| *10* | 7 | 34 | 4 | 7 | **13.0** | **5.9** | **ND** |
| *32* | 4 | 9 | 3 | 31 | **12.0** | **6.1** | **ND** |

**Supplementary Table 5.** *therascreen*® MGMT Kit methylation percentages of discrepant cases when using 5% (cohort 1) and 7% (cohort 2) MGMT promoter (*MGMTp*) methylation as the cutoff with the EpiDirect® MGMT Assay.

---

```r
library(readxl)
library(pROC)

# === STEP 1: Read Excel file ===
data <- read_excel("your_file.xlsx")

# === STEP 2: Extract Methods ===
method1 <- data[[1]]      # Column A: Method 1
methods <- data[, 2:5]    # Columns B–E: Methods 2–5

# === STEP 3: Reference groups from Method 1 (cutoff=10) ===
group1 <- ifelse(method1 < 10, 0, 1)  # 0=Group 1, 1=Group 2 for ROC

# === STEP 4: Function to find ALL optimal ROC cutoffs ===
find_roc_cutoff <- function(method_values) {
  roc_obj <- roc(group1, method_values)

 # Get all optimal cutoffs according to Youden index
  coords_best <- coords(
    roc_obj,
    "best",
    ret = c("threshold", "sensitivity", "specificity"),
    best.method = "youden",
    transpose = FALSE
  )

  # Convert to data frame for easy handling
  best_df <- data.frame(coords_best)

  # Add AUC for context
  best_df$AUC <- as.numeric(auc(roc_obj))

  return(best_df)
}

# === STEP 5: Apply to all methods ===
results <- lapply(methods, find_roc_cutoff)

# === STEP 6: Display results ===
cat("ROC-Optimal Cutoffs vs Method 1 (cutoff=10.00):\n")
for (i in seq_along(results)) {
  cat("\nMethod", i + 1, "\n")
  print(results[[i]])
}

# === OPTIONAL: Plot ROC curves ===
par(mfrow = c(2, 2))
```

```
for (i in 1:ncol(methods)) {
  roc_obj <- roc(group1, methods[[i]])
  plot(roc_obj, main = paste("ROC Curve - Method", i + 1))
  abline(a = 0, b = 1, lty = 2, col = "gray")
```

---

**Supplementary Data 1.** Script used in RStudio for ROC analysis with Youden's Index to find the cutoff for the EpiDirect® MGMT Assay corresponding to a 10% cutoff value used with the therascreen® MGMT Kit.

```
library(readxl)
library(pROC)

# === STEP 1: Read Excel file ===
data <- read_excel(file.choose())
New names:
• `` -> `...6`
• `` -> `...7`
• `` -> `...8`
• `` -> `...9`

# === STEP 2: Extract Methods ===
method1 <- data[[1]] # Column A: Method 1
methods <- data[, 2:5] # Columns B-E: Methods 2-5

# === STEP 3: Reference groups from Method 1 (cutoff = 10) ===
# 0 = below 10
# 1 = 10 or above
group1 <- ifelse(method1 < 10, 0, 1)


# === STEP 4: Function to perform ROC analysis using Youden's Index ===
 # Sensitivity and specificity are weighted equally

 find_roc_cutoff <- function(method_values) {
    # Create ROC curve
    roc_obj <- roc(
       response = group1,
       predictor = method_values,
       levels = c(0, 1),
       direction = "<"
    )
    # Extract all possible threshold points
    coords_all <- coords(
       roc_obj,
       x = "all",
       ret = c("threshold", "sensitivity", "specificity"),
       transpose = FALSE
    )
    coords_df <- data.frame(coords_all)
    # Calculate standard Youden Index:
    # J = Sensitivity + Specificity - 1
    coords_df$youden_index <-
       coords_df$sensitivity + coords_df$specificity - 1
    # Rank cutoffs from highest to lowest Youden Index
    coords_df <- coords_df[
       order(-coords_df$youden_index),
    ]
    # Add AUC for context
    coords_df$AUC <- as.numeric(auc(roc_obj))
    # Return top 10 cutoff points
    head(coords_df, 10)
 }


 # === STEP 5: Apply ROC analysis to Methods 2-5 ===
 results <- lapply(methods, find_roc_cutoff)


 # === STEP 6: Display results ===
 cat(
    "Top 10 ROC Cutoffs Ranked by Youden Index",
    "(Sensitivity and Specificity Weighted Equally)",
    "vs Method 1 (cutoff = 10.00):\n"
```

```
)
Top 10 ROC Cutoffs Ranked by Youden Index (Sensitivity and Specificity Weighted Equally) vs Method 1 (cutoff = 10.00):

for (i in seq_along(results)) {
  cat("\nMethod", i + 1, "\n")
  print(results[[i]])
}

Method 2
   threshold sensitivity specificity youden_index       AUC
30  6.626166   0.8333333   1.0000000    0.8333333 0.9487179
29  5.648911   0.8333333   0.9615385    0.7948718 0.9487179
31  6.965029   0.7777778   1.0000000    0.7777778 0.9487179
28  4.024557   0.8333333   0.9230769    0.7564103 0.9487179
32  7.362196   0.7222222   1.0000000    0.7222222 0.9487179
27  3.126825   0.8333333   0.8846154    0.7179487 0.9487179
24  2.720183   0.8888889   0.8076923    0.6965812 0.9487179
26  2.982968   0.8333333   0.8461538    0.6794872 0.9487179
33  7.655671   0.6666667   1.0000000    0.6666667 0.9487179
23  2.469352   0.8888889   0.7692308    0.6581197 0.9487179

Method 3
   threshold sensitivity specificity youden_index       AUC
30  8.059531   0.8333333   1.0000000    0.8333333 0.9423077
29  6.274768   0.8333333   0.9615385    0.7948718 0.9423077
31  9.026957   0.7777778   1.0000000    0.7777778 0.9423077
28  4.916133   0.8333333   0.9230769    0.7564103 0.9423077
32  9.713925   0.7222222   1.0000000    0.7222222 0.9423077
27  4.550957   0.8333333   0.8846154    0.7179487 0.9423077
24  3.729568   0.8888889   0.8076923    0.6965812 0.9423077
26  4.417481   0.8333333   0.8461538    0.6794872 0.9423077
33 10.324704   0.6666667   1.0000000    0.6666667 0.9423077
23  3.332132   0.8888889   0.7692308    0.6581197 0.9423077

Method 4
   threshold sensitivity specificity youden_index       AUC
30  8.151787   0.8333333   1.0000000    0.8333333 0.9679487
27  6.634693   0.8888889   0.9230769    0.8119658 0.9679487
29  7.455003   0.8333333   0.9615385    0.7948718 0.9679487
31  9.004020   0.7777778   1.0000000    0.7777778 0.9679487
26  6.060027   0.8888889   0.8846154    0.7735043 0.9679487
28  7.049859   0.8333333   0.9230769    0.7564103 0.9679487
23  4.580019   0.9444444   0.8076923    0.7521368 0.9679487
25  5.175428   0.8888889   0.8461538    0.7350427 0.9679487
32 10.171740   0.7222222   1.0000000    0.7222222 0.9679487
22  4.346550   0.9444444   0.7692308    0.7136752 0.9679487

Method 5
   threshold sensitivity specificity youden_index      AUC
30  5.921511   0.8333333   1.0000000    0.8333333 0.957265
29  5.713349   0.8333333   0.9615385    0.7948718 0.957265
31  6.778379   0.7777778   1.0000000    0.7777778 0.957265
28  5.357696   0.8333333   0.9230769    0.7564103 0.957265
23  3.179670   0.9444444   0.8076923    0.7521368 0.957265
25  4.465325   0.8888889   0.8461538    0.7350427 0.957265
32  8.306224   0.7222222   1.0000000    0.7222222 0.957265
27  5.132194   0.8333333   0.8846154    0.7179487 0.957265
22  3.150117   0.9444444   0.7692308    0.7136752 0.957265
24  3.782127   0.8888889   0.8076923    0.6965812 0.957265
```

**Supplementary Data 2.** Script used in RStudio for ROC analysis with Youden's Index to find the cutoff for the EpiDirect® MGMT Assay corresponding to the one used with the therascreen® MGMT Kit when optimizing for an equal positive percent agreement (PPA) and negative percentage agreement (NPA).
